# Effect of mRNA vaccination on pulmonary sequelae after mild COVID-19

**DOI:** 10.1101/2023.12.03.23299330

**Authors:** D Gagiannis, C Hackenbroch, F Zech, F Kirchhoff, W Bloch, K Junghans, K Steinestel

**Affiliations:** Department of Pulmonology, Bundeswehrkrankenhaus Ulm, 89081 Ulm, Oberer Eselsberg 40, Germany; Department of Radiology, Bundeswehrkrankenhaus Ulm, 89081 Ulm, Oberer Eselsberg 40, Germany; Department of Radiology, Ulm University Medical Center, 89081 Ulm, Albert-Einstein-Allee 23, Germany; Institute of Molecular Virology, Ulm University Medical Center, 89081 Ulm, Meyerhofstrasse 1, Germany; Department of Molecular and Cellular Sport Medicine, German Sport University Cologne, 50933 Cologne, Am Sportpark Müngersdorf 6, Germany; Institute of Pathology and Molecular Pathology, Bundeswehrkrankenhaus Ulm, 89081 Ulm, Oberer Eselsberg 40, Germany

**Keywords:** COVID-19, SARS-CoV-2, PASC, lung pathology, longCOVID, postCOVID, vaccination

## Abstract

**Background:** Previous studies indicate a protective role for SARS-CoV-2 vaccination against development of pulmonary post-acute sequelae of COVID (PASC). We compared clinical, imaging, histopathology and ultrastructural features of pulmonary PASC with and without prior vaccination in a consecutive cohort of 54 unvaccinated, 17 partially vaccinated and 28 fully vaccinated patients who presented with dyspnea on exertion after mild COVID-19 (without hospitalization).

**Methods:** Patients underwent full clinical evaluation including autoantibody (ANA/ENA) serology, high-resolution computed tomography (HRCT), bronchioloalveolar lavage fluid (BAL) analysis and transbronchial biopsy followed by histopathological and ultrastructural analysis and SARS-CoV-2 immunohistochemistry.

**Results:** While vaccinated patients were younger (p=0.0056), included more active smokers (p=0.0135) and a longer interval since infection (35 vs. 17 weeks, p=0.0002), dyspnea on exertion and impaired lung function were not different between vaccinated and unvaccinated patients. Ground glass opacities in HRCT and centrilobular fibrosis were more frequent in unvaccinated patients (p=0.0154 and p=0.0353), but presence of autoantibodies, BAL lymphocytosis and bronchiolitis were common findings in all groups. While vaccination against SARS-CoV-2 is associated with a longer time span between infection and consultation along with a reduced frequency of ground glass opacities and centrilobular fibrosis, impaired lung function, bronchiolitis and presence of autoantibodies are comparable between vaccinated and unvaccinated patients. Residual virus was not detected in lung tissue in all but 1 patient.

**Conclusion:** While differences between the investigated groups with regard to age, smoking status and SARS-CoV-2 variants have to be taken into account, a proposed protective role of SARS-CoV-2 vaccination against pulmonary PASC is so far not fully explained by clinical and histopathology findings.

**KEY MESSAGES:** The role of SARS-CoV-2 vaccination in the protection against pulmonary post-acute sequelae of COVID-19 (PASC) is unclear. Using a multidimensional approach integrating clinical, serological, imaging and histopathology data as well as ultrastructural analyses, we show here that previous vaccination has no impact on lung function, bronchiolitis or the detection of autoantibodies or residual virus in a previously healthy cohort of 99 PASC patients after mild COVID-19. While a higher frequency of ground glass opacities in unvaccinated patients might be due to the longer interval between infection and consultation, the observed fibrotic remodeling should prompt further investigation of a possible pro-fibrotic role of SARS-CoV-2 infection in the lung.

## INTRODUCTION

Following SARS-CoV-2 infection, up to 15% of non-hospitalized patients develop post-infectious sequelae of COVID-19 (PASC) [1-3]. Lung manifestations of PASC include shortness of breath, chest pain and imaging abnormalities such as ground glass opacities (GGO) and subpleural bands [4]. Evaluation of PASC can be hampered by preexisting lung disease or severe COVID-19 with scarring or post-ventilaton change [5]. In a a prospective cohort of 51 previously healthy pulmonary PASC patients after mild COVID-19, we have previously shown pulmonary function impairment in a subset of patients, including a reduction in maximal expiratory flow at 50% of FVC (MEF50) and a reduction in FEV1 together with CD4+T cell-mediated bronchiolitis, indicating obstructive change [6, 7]. To our best knowledge, this was the first study to use verifiable tissue-based criteria to assess lung involvement in PASC in previously healthy patients after mild COVID-19. In several large cohort studies, a reduction in both frequency and symptom severity of pulmonary PASC upon mRNA vaccination against SARS-CoV-2 has been described [8-10]. However, since most of these reports are based on self-reported symptoms or electronic medical records, the aim of the present study was to evaluate the effect of previous vaccination on defined clinical, imaging and histopathological characteristics of pulmonary PASC.

## METHODS

### Patients and diagnostic work-up

We consecutively included n=99 patients with a history of RT-PCR-confirmed SARS-CoV-2 infection and persistent pulmonary symptoms in the present study. 54 patients were unvaccinated, 17 patients were partially vaccinated (1 to 2 doses) and 28 patients were fully vaccinated (3 or more doses) at the time of SARS-CoV-2 infection. All vaccinations were performed using the BNT126b2 mRNA vaccine (BionTech/Pfizer). The baseline diagnostic workup included clinical history including current and previous medication and allergies, physical examination and spiroergometry. ANA/ANCA/ENA screening (serology) was performed as previously described [11]. After multidisciplinary discussion, bronchoscopy with transbronchial biopsy for the assessment of possible PASC-ILD was performed in all patients that formed the definite cohort. Patients had given written informed consent to routine diagnostic procedures (serology, bronchoscopy, imaging) as well as to the scientific use of data and tissue samples in the present study. This project was approved by the local ethics committee of the University of Ulm (ref. no. 129-20) and conducted in accordance with the Declaration of Helsinki.

### Imaging

Imaging of the lung was performed on a 3rd generation DECT Scanner (Siemens Somatom Force, Siemens Healthineers, Forchheim, Germany). A non-contrast multislice spiral CT in fulldose technique (Qual. Ref. kV: Sn 100, Qual. Ref. mAs: 600, ref. CTDI 1.06 mGy) with a collimation of 192 × 0.6 mm and using a tin prefilter at 100 resp. 150 kV was obtained. Rotation time 0.25 sec, scan time 1.25 sec. Examination was performed in inspiration. Reconstruction in bone, soft tissue and lung window.

### Histology and SARS-CoV-2 detection

Lung tissue specimens were obtained as transbronchial biopsies and stained with haematoxylin-eosin (HE), Elastica-van-Gieson (EvG) and Masson-Goldner (MG). Slides were reviewed with special emphasis on possible post-viral change (alveolitis, peribronchiolitis, airway smooth muscle hypertrophy, goblet cell hyperplasia, airway squamous epithelial metaplasia, and fibrosis)[12]. We performed immunohistochemistry for Spike and Nucleocapsid proteins of SARS-CoV-2 using the following antibodies: anti-SARS-CoV-2 spike antibody, clone 1A9, mouse monoclonal, 1:400 (GeneTex, Irvine, CA, USA); anti-SARS-CoV-2 nucleocapsid antibody, rabbit monoclonal, 1:20000 (SinoBiological, Bejing, China). Tissue-based RT-PCR in a subset of cases was performed as previously described [7].

### Electron microscopy

Lung tissue was immersion-fixed with 4% paraformaldehyde in 0.1M PBS, pH 7.4. After several washing steps in 0.1M PBS, tissue was osmicated with 1% OsO4 in 0.1 M cacodylate and dehydrated in increasing ethanol concentrations. Epon infiltration and flat embedding were performed following standard procedures. Methylene blue was used to stain semithin sections of 0.5 μm. Seventy to ninety-nanometer-thick sections were cut with an Ultracut UCT ultramicrotome (Fa. Reichert) and stained with 1% aqueous uranylic acetate and lead citrate. Samples were studied with a Zeiss EM 109 electron microscope (Fa. Zeiss) coupled to a TRS USB (2048x2048, v.596.0/466.0) camera system with ImageSP ver.1.2.6.11 (x64) software.

### BAL analysis and legendplex immunoassay

The Legendplex ELISA was performed according to the manufacturer’s instructions. In brief, 25 μl of the collected BAL was centrifuged (4000 rpm, 5 min) and incubated for 2 h at room temperature with antibody-coated beads, followed by washing and incubation with the detection antibodies. After incubation with the staining reagent, the beads were analyzed in a high-throughput sampler via flow cytometry (FACSCanto II, BD Biosciences). Absolute quantification was performed using a standard and the Bio-legend Legendplex v8.0 software.

### Statistical Methods

Descriptive statistical methods were used to summarize the data (**Table 1**). Medians and interquartile ranges were used to announce results. Absolute numbers and percentages were employed to represent categorial variables. Student’s t-test/ANOVA with multiple comparison post-test was used for the comparison of continuous variables, while Chi-Square-Test/Fisher’s test was used for categorial variables. All statistical analyses were conducted using GraphPad PRISM 10.1.0 (GraphPad Software Inc., San Diego, CA, USA). A p-value <0.05 was regarded as statistically significant.

**Table 1.** Clinical characteristics, lung function and imaging/histopathology results in PASC patients stratified according to vaccination status.

| Characteristics | Unvaccinated<br>(n=54) | partially vaccinated<br>(n=17) | fully vaccinated<br>(n=28) | p-value |
| --- | --- | --- | --- | --- |
| **Age (years; median, range)** | 41 (18-81) | 39 (24-61) | 29 (19-73) | **0.0056\*\*** |
| **Gender, n (%)** |  |  |  |  |
| Female | 19 (35) | 8 (47) | 17 (61) | 0.0806 |
| Male | 35 (65) | 9 (53) | 11 (39) |  |
| **Smoking status, n (%)** |  |  |  |  |
| Current smoker | 5 (9) | 5 (29) | 11 (39) | **0.0135\*** |
| Former smoker | 12 (22) | 1 (6) | 4 (14) |  |
| Never smoker | 37 (69) | 11 (65) | 13 (46) |  |
| **Weeks since SARS-CoV-2, median (range)** | 17 (2-56) | 22 (13-100) | 35 (13-91) | **0.0002\*\*\*** |
| **Presence of autoantibodies<sup>1</sup>, n (%)** | 20 (37) | 10 (59) | 13 (46) | 0.2579 |
| **FVC, % pred, median (IQR)** | 93 (56-120) | 90 (68-110) | 91 (34-116) | 0.3549 |
| **TLC, % pred, median (IQR)** | 103 (63-148) | 96 (81-149) | 104 (57-142) | 0.9556 |
| **FEV1, % pred, median (IQR)** | 93 (63-123) | 85 (65-112) | 91 (40-116) | 0.4239 |
| **MEF50, % pred, median (IQR)** | 91 (45-175) | 79 (49-156) | 79 (43-127) | 0.378 |
| **D<sub>LCO</sub>, % pred, median (IQR)** | 94 (53-210) | 89 (69-124) | 96 (72-123) | 0.564 |
| **VO2/HF, median (IQR)** | 14 (3-37) | 12.9 (9.1-15.4) | 10.9 (6.8-19.3) | **0.034\*** |
| **Low FVC <80% pred, n (%)** | 7 (13) | 3 (18) | 6 (21) | 0.612 |
| **Low TLC <80% pred, n (%)** | 4 (7) | 0 | 1 (4) | 0.5753 |
| **Low FEV1 <80% pred, n (%)** | 10 (19) | 4 (24) | 6 (21) | 0.3224 |
| **Low MEF50 <80% pred, n (%)** | 20 (37) | 9 (53) | 14 (50) | 0.3612 |
| **Low D<sub>LCO</sub> <70% pred, n (%)** | 4 (7) | 2 (12) | 0 | 0.1961 |
| **BAL** |  |  |  |  |
| Lymphocyte count, median (IQR) | 12,5 (1,8-59,6) | 13,65 (5,8-40,1) | 20,5 (2,8-58) | 0.6623 |
| **Imaging, n (%)** |  |  |  |  |
| Ground glass opacities | 9 (17) | 1 (6) | 0 | **0.0394\*** |
| Reticulation | 5 (9) | 2 (12) | 4 (14) | 0.086 |
| Noduli | 0 | 2 (12) | 1 (4) | **0.0495\*** |
| **Histopathology, n (%)** |  |  |  |  |
| Organizing pneumonia | 1 (2) | 2 (12) | 1 (4) | 0.2128 |
| Lymphoid follicles | 14 (26) | 3 (18) | 3 (11) | 0.3029 |
| Fibrosis | 13 (24) | 0 | 3 (11) | **0.0353\*** |
| **Interstitial Fibrosis (TEM), n (%)** |  |  |  |  |
| light | 0 | 2 (12) | 9 (32) | **0.0021\*\*** |
| severe | 13 (24) | 1 (6) | 9 (32) |  |

<sup>1</sup>ANA titer ≥ 1:320 and/or positive IB (Scl-70, PCNA, PM-Scl, dsDNA, SS-B, Histone) or RF > 20 U/ml; FVC: forced vital capacity; IQR, interquartile range; TLC: total lung capacity; FEV1: forced expiratory volume in 1 s; MEF50: mean expiratory flow at 50% of FVC; D<sub>LCO</sub>: diffusing capacity of the lung for carbon monoxide; VO2/HF: oxygen pulse; BAL: broncho-alveolar lavage; TEM, transmission electron microscopy; \*p<0.05; \*\*p<0.005; \*\*\*p<0.001

## RESULTS

### Diagnostic workup, clinical data and results from lung function tests

The diagnostic work-up of suspected PASC-ILD patients who presented to our outpatient clinic at the Bundeswehrkrankenhaus (army hospital) Ulm is summarized in **Fig. 1 A**, while **Tab. 1** and **Fig. 1 C** summarize clinical, serological, imaging and histopathology data for the complete cohort. 51/54 patients from the unvaccinated cohort have been published previously [7]. No patient had been hospitalized for COVID-19, 61/99 patients were soldiers (61.6%). Vaccinated patients were significantly younger (p=0.0056), active smokers (p=0.0135) and seeked consultation after a longer time span following acute SARS-CoV-2 infection (p=0.0002, **Fig. 1 B**). Of note, while the recruitment period for unvaccinated patients was 11/2020-04/2021, the recruitment period for vaccinated patients was 10/2021-04/2023, implying a different spectrum of SARS-CoV-2 viral variants. The most frequent pathological result from pulmonary function test were impaired MEF50 in 37%, 53% and 50% and and FEV1 (≤80%) in 19%, 24% and 21% of patients in the vaccinated, partially vaccinated and fully vaccinated groups, respectively (p>0.05). Median oxygen pulse (VO_2_/HF) was significantly lower in the fully vaccinated compared to the unvaccinated group (p=0.034). With respect other lung function tests, there was no significant difference between the groups according to vaccination status (**Fig. 2 A**). Furthermore, there was no significant association between the time since COVID and the number of lymphocytes in BAL or oxygen pulse in any of the investigated groups, although there was a borderline correlation between oxygen pulse and time since COVID in unvaccinated patients (p=0.0605, **Fig. 2 B**)

**Figure 1.**
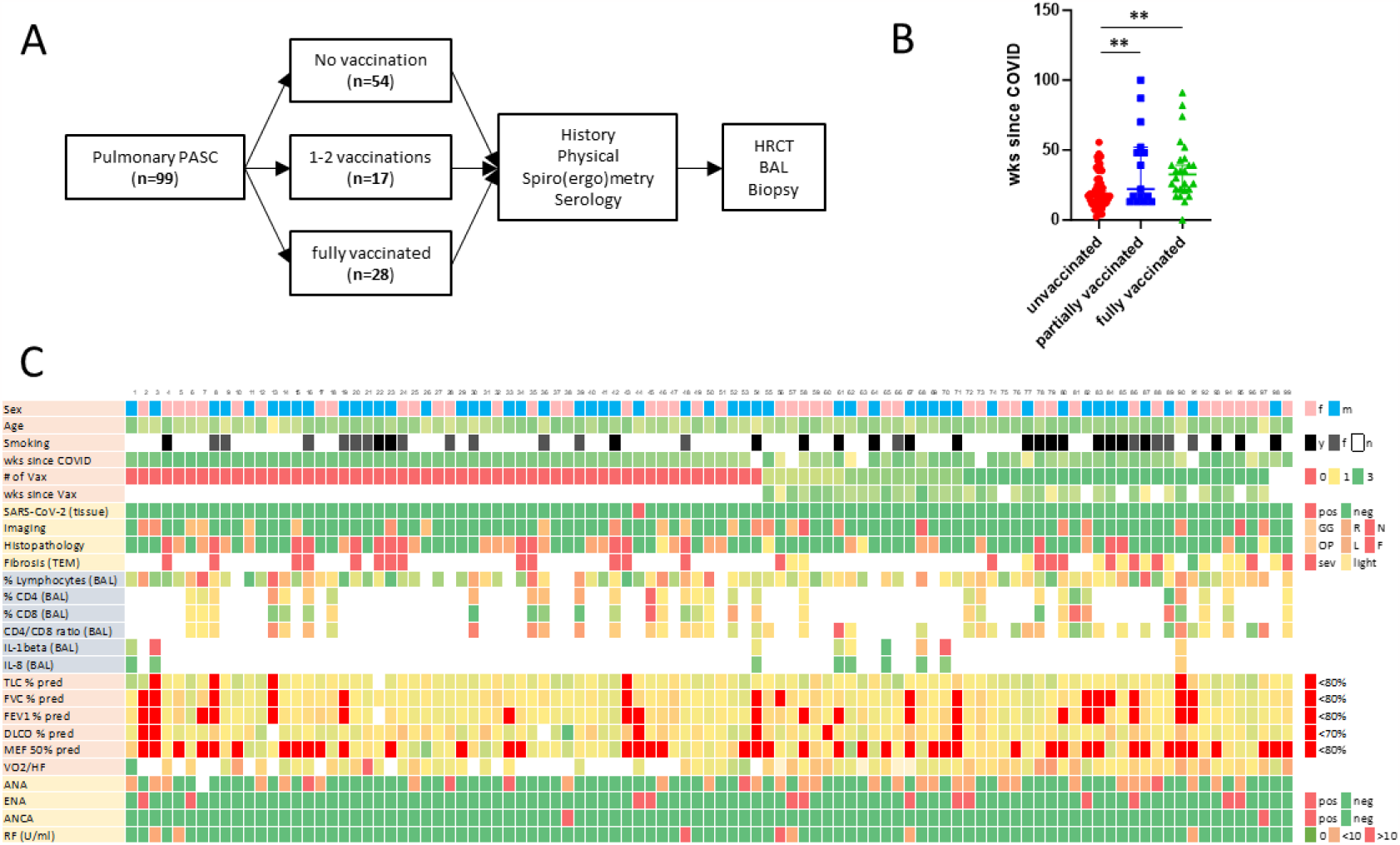
Characterization and diagnostic work-up of PASC patients. **A**, Flow diagram for diagnostic work-up of suspected PASC-ILD patients. Lung biopsies from 99 PASC patients were finally included in histopathological assessment. **B**, interval between infection and clinical presentation. *\*\*p<0.05* **C**, heatmap stratified according vaccination status displaying clinical data and the results from tissue-based SARS-CoV-2 detection, imaging, histopathology, BAL analysis, lung function tests and autoantibody screening.

**Figure 2.**
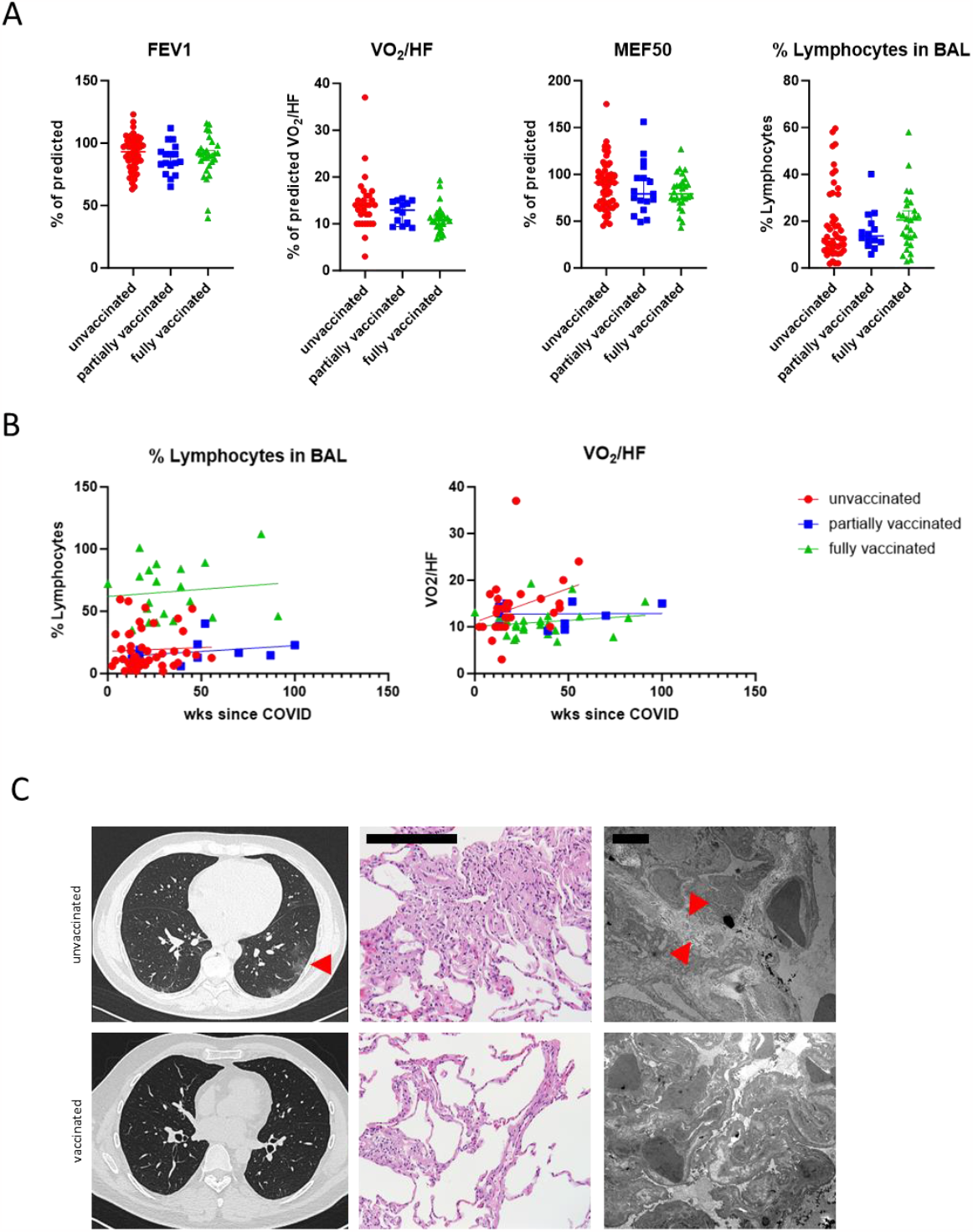
Lung function tests, BAL analyses and representative images from histopathology and HRCT. **A**, No significant difference between FEV1, oxygen pulse, MEF50 and BAL lymphocyte count according to vaccination status. **B**, No significant correlation between oxygen pulse or BAL lymphocyte count and time since infection, irrespective of vaccination status. **C**, Representative HRCT images showing ground glass opacities (arrowhead) in an unvaccinated compared to a vaccinated PASC patient vs. (left panel). Representative microphotographs of H.E.-stained lung tissue samples from an unvaccinated PASC patient showing interstitial lymphocytosis and discrete centrilobular fibrosis (center panel). *Scale bar, 200μm*. Ultrastructural analyses show interstitial collagen deposition (arrowhead) in an unvaccinated PASC patient.

### Serology and imaging

Autoantibodies (serology; ANA titer ≥ 1:320 and/or positive immunoblot for Scl-70, PCNA, PM-Scl, dsDNA, SS-B, Histone) were detected in 37, 59 and 56% of unvaccinated, partially vaccinated and fully vaccinated PASC patients. There was no difference in the frequency of autoantibodies, the inflammatory infiltrate in BAL samples (both p>0.05) or BAL cytokine profile between vaccinated and unvaccinated patients. In HRCT imaging, ground glass opacities (GGO) were significantly more frequent in the unvaccinated compared to the vaccinated groups (p=0.0154) (**Fig. 2 C**), while noduli were observed more often in vaccinated patients (p=0.0495).

### Histopathologic findings and tissue-based SARS-CoV-2 detection

Histopathologic evaluation of transbronchial biopsies from PASC patients showed increased peribronchiolar and interstitial lymphocytosis and alveolar fibrin deposition irrespective of vaccination status (**Fig. 2 C**). Organizing pneumonia was rare in all groups (1 unvaccinated, 2 partially vaccinated and 1 unvaccinated patient), while lymphoid follicles were more frequent in unvaccinated patients without reaching statistical significance. Of note, centrilobular fibrosis was significantly increased in unvaccinated compared to vaccinated patients as assessed by Masson-Goldner staining/ light microscopy (p=0.0353). This could be confirmed by ultrastructural analyses (transmission electron microscopy), where all investigated cases in the unvaccinated cohort showed marked collagen deposition (p=0.0021). Immunohistochemistry for SARS-CoV-2 N and S proteins was negative in all tissue samples but one previously published case in the unvaccinated cohort was positive for E gene in RT-PCR which could be confirmed by SARS-CoV-2 S FISH analysis of the alveolar epithelium [7].

## DISCUSSION

We report on histopathological, serological, clinical and imaging characteristics of 99 pulmonary PASC patients with and without previous vaccination from the postCOVID outpatient clinic of the Bundeswehrkrankenhaus (army hospital) Ulm to assess whether previous vaccination has an impact on verifiable and tissue-based criteria of PASC severity.

In comparison to most published PASC cohorts, our patient cohort was younger and had less preexisting medical conditions, both possibly due to the high proportion of active soldiers in the study and reflecting data from other studies involving military personnel [13]. All patients presented with dyspnea on exertion irrespective of vaccination status, but the time span between infection and presentation was significantly longer in vaccinated patients. While this might reflect less symptomatic disease, it has to be taken into account that while the unvaccinated cohort was in most part recruited during the SARS-CoV2 delta wave, infections in the vaccinated cohort occurred during spread of the Omicron variant. Moreover, media coverage in 2020/21 might have contributed to a higher level of alertedness in affected patients which resulted in earlier consultation.

With respect to lung function, there were no significant differences between vaccinated and unvaccinated patients. In line with previous results from comparable cohorts, reduced FEV1 and MEF50 were the most prevalent finding [7, 14]. While there was a significant lower oxygen pulse in fully vaccinated compared to unvaccinated patients, we would ascribe this finding to the higher number of active smokers in this group. Of note, there was a strong trend towards higher oxygen pulse in unvaccinated patients with increasing interval since infection that could not be observed in the vaccinated groups. The relevance of this finding is unclear, but it might be speculated that this trend is due to the shorter time interval since infection and more nuanced differences in VO_2_/HF among the unvaccinated cohort.

While we would question the higher frequency of nodular changes in HRCT in the vaccinated group due to the low absolute number of observations, imaging showed significantly higher frequency of GGOs in unvaccinated PASC patients. This might be due to alveolar fibrin deposition which we have previously reported as an almost ubiquitous finding in lung histopathology of both acute COVID-19 and PASC [7, 15]. It is well conceiveable that the shorter time span between infection and imaging in the unvaccinated group contributed to that finding, with temporal resolution of alveolar fibrin in the other groups.

Among all patients, (peri)bronchiolar chronic inflammation and alveolar fibrin deposition were common findings in histopathology. Organizing pneumonia as another possible correlate for GGOs was a rare finding in histopatholoy of lung tissue samples among all groups, while fibrotic change was more frequent among unvaccinated patients. This finding, which could be confirmed by electron microscopy, is of relevance since we have previously demonstrated elevated levels of IF-1β in BAL fluid, ultrastructural fibrosis and accumulation of pro-fibrotic macrophages in PASC [7]. Moreover, we and others have shown that SARS-CoV-2 has the ability to induce lung fibrosis [15, 16]. While the impact of severe COVID-19 pneumonia or invasive ventilation on fibrotic remodeling is well-demonstrated, further research is warranted to understand possible pro-fibrotic signaling during and after a mild course of COVID-19 [17].

With respect to the pathophysiology of pulmonary PASC, autoimmunity and viral persistence have been discussed as possible contributors [18, 19]. While we detected autoantibodies in a large fraction of PASC patients, their presence did neither correlate with vaccination status nor with the severity of symptoms, why we conclude that if PASC is linked to the presence of autoantibodies, vaccination is unlikely to prevent their formation. Because the presence of autoantibodies is associated with severe course of acute COVID-19 and has been discussed as a hallmark of extrafollicular B cell activation upon contact with SARS-CoV-2 however, we still think that the role of autoimmunity in both acute COVID-19 and PASC should be further investigated including research on lung tissue samples [20, 21]. With respect to possible viral reservoirs, there were no detectable viral proteins in lung tissue samples from all 99 investigated patients, irresprective of vaccination status. In one patient, viral mRNA could be detected by RT-PCR. We conclude persistence of virus/viral particles is not necessary for PASC, in line with the situation in acute COVID-19, where organization and long-term outcomes occur independent from the presence of virus [15].

There are clear limitations to our study. First, these are real-world cohorts that bear significant differences with respect to age, smoking status and viral variants between the vaccinated and unvaccinated cohorts. Still, since symptom severity and viral variants would have favoured the vaccinated cohort, we think that our findings contribute important evidence for the assessment of a possible role of vaccination in the prevention of postCOVID. Second, since a significant proportion of investigated patients are active soldiers, the meaning for the general population might be limited. With respect to that, we think that young age and the absence of relevant pulmonary pathology is a unique opportunity to investigate the pathology of PASC and the role of vaccination in that setting. Third, the value of transbronchial biopsies for the assemssent of interstitial lung disease is limited, however most studies that combine lung function tests with analyses on peripheral blood lack such opportunity for multidimensional characterization of a condition that affects lung tissue.

Taken together, our data shows impairment of lung function (MEF50/FEV1), presence of autoantibodies and BAL lymphocytosis along with centrilobular inflammation in PASC patients irrespective of vaccination status. While GGO and centrilobular fibrosis seem to occur more frequently in unvaccinated patients, this difference might as well be due to the longer time interval between infection and consultation in vaccinated patients. While our data supports research effort with the respect of inflammasome and autoimmunity in the general pathogenesis of pulmonary PASC, it does not support a protective role of mRNA vaccination against the development of bronchiolitis and obstruction as possible contributors to the clinical presentation of PASC.

## Data Availability

All data produced in the present study are available upon reasonable request to the authors

## ACKNOWLEDGEMENTS

The authors would like to thank all patients for their consent to the use of data and images in the present study.

## AUTHOR CONTRIBUTIONS

Study concept: DG and KS. Data collection: DG, CH, FZ, FK, KJ, KS. Sample collection: DG, CH, KS. Initial draft of manuscript: KS. Critical revision and approval of final version: all authors.

